# Dissecting the genetic etiology of intestinal obstruction: mendelian randomization identifies potential therapeutic targets

**DOI:** 10.1101/2025.02.19.25322532

**Authors:** Zhao Haodong, Cai Baoping, Chen Jiongjiong, Chen Jia, Liu hui, Wang Yu, Ding bocheng

## Abstract

Intestinal obstruction, a prevalent and serious condition, necessitates deeper understanding of its genetic architecture. This study rigorously employed two-sample Mendelian randomization to dissect the causal influence of gene expression on intestinal obstruction risk, leveraging comprehensive summary-level data from eQTLGen and FinnGen GWAS. To enhance causal inference, Summary-data-based Mendelian Randomization was integrated, utilizing GTEx eQTL data to specifically assess tissue-relevant gene expression. Our multi-pronged analyses provide compelling genetic evidence supporting causal roles for genetically predicted expression of *CHRNB2* and *MIAT* in intestinal obstruction. Specifically, increased genetically proxied *CHRNB2* expression was associated with a protective effect, while higher genetically proxied *MIAT* expression suggested an elevated susceptibility to intestinal obstruction. Colocalization analysis, alongside HEIDI heterogeneity testing within the SMR framework, further bolstered the robustness of these findings by distinguishing causality from linkage disequilibrium. These convergent results offer novel mechanistic insights into intestinal obstruction pathogenesis, positioning *CHRNB2* and *MIAT* as promising therapeutic targets for both prevention and treatment strategies. Future research is crucial to validate these findings across diverse ancestries and to fully elucidate the intricate biological mechanisms underpinning these gene-disease associations.

## Introduction

Intestinal obstruction represents a significant clinical challenge, affecting millions worldwide and posing a potentially life-threatening threat that necessitates prompt medical or surgical intervention. It arises from a variety of causes, including postoperative adhesions, hernias, malignancies, and inflammatory disorders such as Crohn’s disease[1]. Despite advances in surgical techniques and critical care management, the pathophysiological mechanisms underlying intestinal obstruction remain poorly understood, and preventive strategies are limited[2]. Recent evidence suggests that genetic factors may contribute to individual susceptibility to intestinal obstruction, yet the causal genes and molecular pathways involved remain largely unknown[3, 4].

Mendelian randomization (MR) has become increasingly recognized as a robust method to infer causal relationships between risk factors and disease outcomes by leveraging genetic variants as instrumental variables[5]. Unlike observational studies, which are often confounded by reverse causation and environmental biases, MR mimics a randomized controlled trial by utilizing genetic variants that are randomly allocated at conception, making it particularly well-suited for investigating complex diseases like intestinal obstruction where traditional randomized controlled trials are often infeasible or unethical[6]. In recent years, expression quantitative trait loci (eQTL) data have been increasingly integrated into MR frameworks to investigate the causal effects of gene expression on disease risk, offering valuable perspectives on gene-disease relationships [7, 8].

To enhance the rigor and reliability of causal inference from MR studies, colocalization analysis is often employed alongside MR to determine whether the same genetic variant drives both gene expression and disease risk, rather than being a result of linkage disequilibrium (LD)[9]. Additionally, Summary-data-based Mendelian Randomization (SMR) combined with the HEIDI test can further distinguish true causal relationships from LD-driven associations[10]. These complementary approaches enhance the robustness of genetic findings and help prioritize candidate genes for further functional validation.

This study aimed to identify potential causal genes for intestinal obstruction by leveraging large-scale eQTL and GWAS data. We conducted a two-sample MR analysis using eQTL data from the eQTLGen consortium and GWAS summary statistics from the FinnGen database. To validate our findings, we performed colocalization analysis using the COLOC method and further applied SMR with HEIDI testing to distinguish true causal effects from LD artifacts. Our study undertakes a comprehensive Mendelian randomization analysis, integrating multiple validation methods, to elucidate the genetic landscape of intestinal obstruction. Our findings are expected to yield novel insights into the pathogenesis of intestinal obstruction, pinpoint potential molecular targets for therapeutic intervention, and pave the way for future research and drug development efforts in this clinically significant condition.

## Methods

### Approval and Data Sources

This study uses publicly available, summary-level GWAS data from the eQTLGen consortium[11], which provides blood-based cis-eQTL data, and FinnGen[12], which contains GWAS data comparing intestinal obstruction patients and controls. GTEx (version 8) [13] eQTL data were used solely for the SMR validation step to assess tissue-specific gene expression relevance. These data were not incorporated into the primary Mendelian randomization (MR) analysis, which relied exclusively on eQTLGen data. The study workflow is illustrated in Fig 1. All data sources have existing approvals from their respective Institutional Review Boards, and all participants provided written informed consent. Therefore, no additional ethical approval was required for this study.

**Fig 1.**
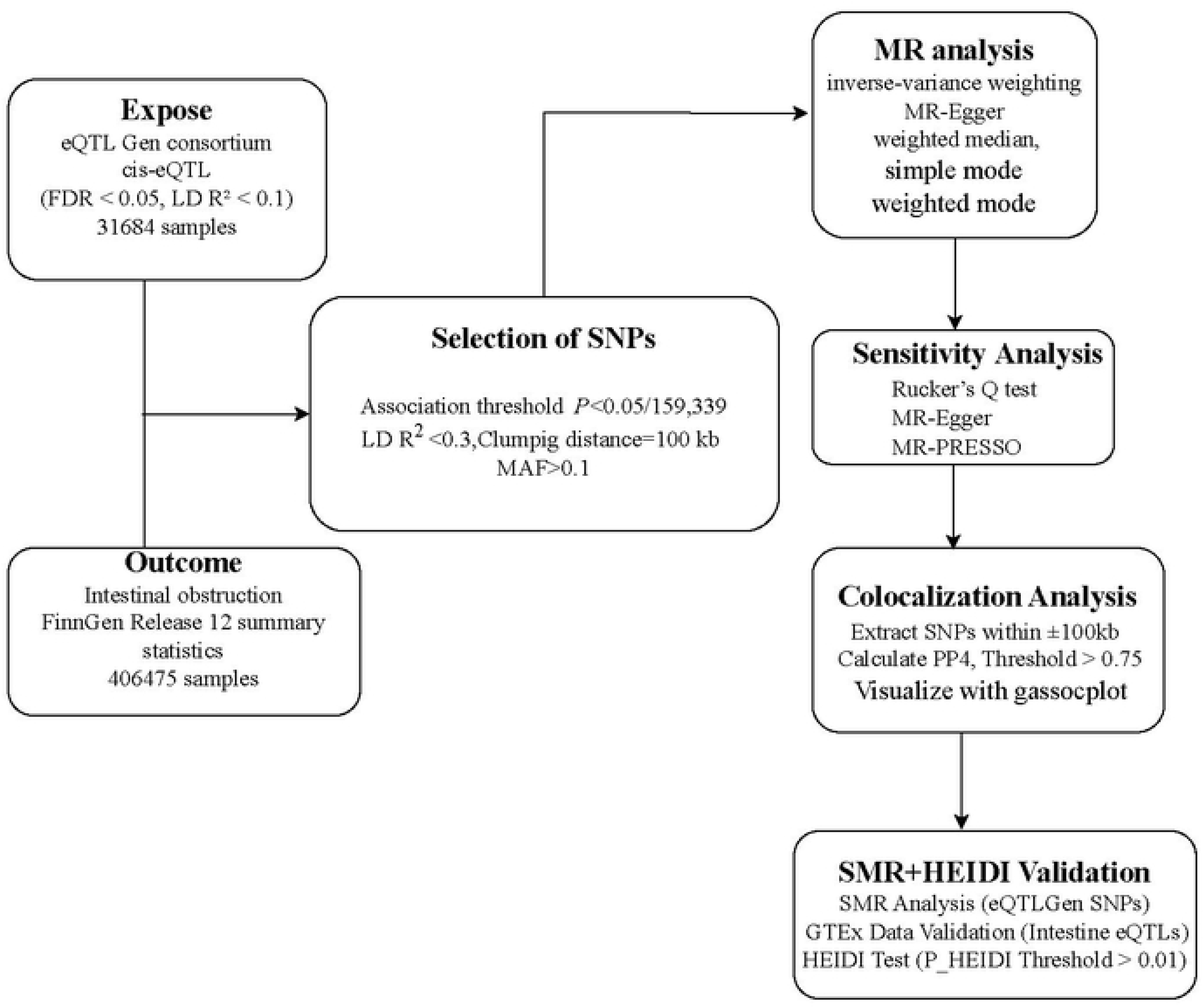
Mendelian Randomization Analysis and Validation Workflow for Genetic Causal Genes of Intestinal Obstruction.

### Genetic Instrument Selection

To select valid eQTL instruments for Mendelian randomization analysis, we applied stringent filtering criteria to ensure statistical robustness and biological relevance. Cis-eQTL variants from the eQTLGen consortium, located within ±100 kb of the target gene, were considered to ensure a direct regulatory effect on gene expression. To control for false positives, we applied false discovery rate (FDR) correction at a threshold of 0.05, retaining only SNPs with statistically significant associations after multiple testing adjustment. To minimize LD confounding, we implemented LD clumping (R² < 0.1, window size = 100 kb) using 1000 Genomes European reference data, ensuring independence between selected SNPs. After applying these criteria, we obtained a set of high-confidence IVs suitable for Mendelian randomization, reducing the risk of bias and improving the reliability of causal inference. The exposure dataset consisted of instrumental variables corresponding to gene expression, selected based on the Inverse Variance Weighted (IVW) method, with the significance threshold determined by Bonferroni correction (*P* < 0.05 / total number of SNPs analyzed). Each gene was analyzed separately for its association with intestinal obstruction risk using summary-level GWAS data from FinnGen as the outcome.

### Mendelian Randomization Analysis

To investigate the causal relationship between gene expression levels and the risk of intestinal obstruction, we conducted two-sample MR. The MR pipeline included instrument selection, LD filtering, harmonization, and causal inference estimation. Instrumental variables from the eQTLGen databases were matched with FinnGen GWAS summary statistics for intestinal obstruction, ensuring consistent effect allele orientation and excluding palindromic SNPs with intermediate allele frequencies. To mitigate confounding due to linkage disequilibrium, we applied LD clumping (R² < 0.3, window size = 100 kb) using reference data from the 1000 Genomes European, depending on the ancestry of the dataset. SNPs with minor allele frequency (MAF) < 1% were excluded. MR analyses were conducted using IVW, MR-Egger regression, and Weighted Median methods to estimate causal effects while accounting for potential pleiotropy.

Sensitivity analyses included heterogeneity testing (Cochran’s Q test) and horizontal pleiotropy assessment (MR-Egger intercept), to evaluate the robustness of findings. Additional outlier detection was performed using MR-PRESSO, and results were visualized using forest plots and scatter plots. The final MR estimates were reported as odds ratios (ORs) with 95% confidence intervals (CIs) and P-values for each target gene.

### Colocalization Analysis

To determine whether the genetic associations identified in MR share a causal variant with intestinal obstruction risk rather than being driven by linkage disequilibrium, we performed colocalization analysis using the COLOC method. For each gene identified in MR, we first determined the associated genomic region using eQTL results and defined a ±100 kb window around the top SNP. Summary-level GWAS data from FinnGen were extracted and harmonized with the exposure dataset to ensure consistency. Colocalization analysis was conducted using the coloc.abf function, which calculates the posterior probability of different colocalization hypotheses: PP4 > 0.75 suggests strong evidence that gene expression and intestinal obstruction share a causal variant. PP3 > PP4 indicates distinct causal variants, suggesting MR results may be influenced by LD rather than true colocalization. Colocalization analysis was visualized using the locuscomparer package.

### SMR Validation Analysis

To validate the findings from Mendelian randomization, we conducted SMR analysis, using eQTL data from eQTLGen and GWAS summary statistics for intestinal obstruction from FinnGen. Additionally, we incorporated tissue-specific eQTL data from GTEx V8, focusing on intestinal tissue, to further assess the relevance of gene expression in the context of intestinal obstruction.

SMR evaluates whether genetic variants associated with gene expression also influence disease risk, providing additional support for a causal relationship between gene expression and intestinal obstruction. For each gene identified in MR, Cis-eQTL SNPs within ±100 kb of the target gene were extracted and matched to the FinnGen GWAS dataset. SMR analysis was then conducted to assess whether genetically predicted gene expression levels were associated with intestinal obstruction risk. The SMR test operates on the principle that if a genetic variant influences both gene expression and disease risk, and this association is mediated through gene expression, the effect sizes from eQTL and GWAS should be proportional. Using a summary-data Mendelian randomization framework, SMR evaluates whether the observed association aligns with this expectation, with the significance of the association determined by the SMR test statistic and corresponding *P*-value (*P*-SMR), which indicates whether the GWAS signal at a given locus is likely mediated through gene expression. To distinguish true colocalization from LD effects, we applied the Heterogeneity in Dependent Instruments (HEIDI) test, which evaluates the heterogeneity of instrument effects across multiple SNPs. If *P*-HEIDI < 0.01, it suggests that the observed association is likely driven by LD rather than a shared causal variant. SMR and HEIDI analyses were conducted for each gene identified as significant in MR, serving as a validation step to confirm that the associations observed in the primary MR analysis were not confounded by LD-related biases.

### Statistical Analysis

All statistical analyses were conducted in R (version 4.4.2) using the Two Sample MR and COLOC packages. SMR analysis was performed using the GCTA-SMR tool. Mendelian randomization estimates were obtained using IVW, MR-Egger, and Weighted Median methods, with significance determined at *P* < 0.05. Colocalization analysis was performed using COLOC, with PP4 > 0.75 considered strong evidence for colocalization. SMR analysis was used as an additional validation tool, with *P*-HEIDI > 0.01 supporting colocalization and *P-* HEIDI < 0.01 suggesting LD confounding. Multiple testing correction was applied using FDR (Benjamini-Hochberg method) where appropriate.

## Results

### Selection of Genetic IVs

We selected genetic IVs from the eQTLGen consortium (Release 2) using predefined filtering criteria (FDR < 0.05, LD R² < 0.1), ensuring independence and statistical robustness. The full list of eQTL variants before MR analysis is detailed in S1 Table. We then performed two-sample MR analysis using 159,339 Cis-eQTL variants to assess their associations with intestinal obstruction based on FinnGen GWAS data. For each gene, IVW analysis was conducted to estimate the effect size and statistical significance of gene expression on intestinal obstruction risk. To account for multiple testing, we applied Bonferroni correction and set the significance threshold at P < 3.14 × 10⁻⁷, a threshold obtained by dividing 0.05 by the number of tests, 159,339(S2 Table). Genes meeting this threshold were selected as final genetic IVs. The selected genes were *ATG12* (*P* = 1.07 × 10⁻⁹, OR = 1.18, 95% CI: 1.12-1.24), *CHRNB2* (*P* = 3.00 × 10⁻⁸, OR = 0.66, 95% CI: 0.57-0.78), *TNFRSF18* (*P* = 4.25 × 10⁻⁸, OR = 0.84, 95% CI: 0.79-0.89), and *MIAT* (*P* = 2.36 × 10⁻⁷, OR = 1.10, 95% CI: 1.06-1.14). The number of IVs per gene varied, with *ATG12* containing the most (23 SNPs) and *CHRNB2* containing the least (6 SNPs). The details of the final selected IVs are summarized in Table 1.

**Table 1.** Final Selected Genetic IVs from eQTLGen for MR Analysis of Intestinal Obstruction.

| GENE | Method | nSNP | $\beta$ | SE | *P* | OR(CI) |
| --- | --- | --- | --- | --- | --- | --- |
| *ATG12* | IVW | 23 | 0.16 | 0.03 | 1.07E-09 | 1.18 ( 1.12-1.24 ) |
| *CHRNA2* | IVW | 6 | -0.41 | 0.07 | 3.00E-08 | 0.66(0.57-0.78) |
| *TNFRSF18* | IVW | 10 | -0.18 | 0.03 | 4.25E-08 | 0.84(0.79-0.89) |
| *MIAT* | IVW | 26 | 0.10 | 0.02 | 2.36E-07 | 1.10(1.06-1.14) |

Abbreviations: IVs = instrumental variables, MR = Mendelian randomization, SNP = single-nucleotide polymorphism, IVW = inverse-variance weighted, SE = standard error, OR = odds ratio, CI = confidence interval.

### MR Analysis Results

To evaluate the potential causal effect of gene expression levels on the risk of intestinal obstruction, we performed a two-sample Mendelian randomization (MR) analysis. Inverse Variance Weighted (IVW) method was employed as the primary analysis, complemented by MR-Egger regression, Weighted Median, Simple Mode, and Weighted Mode methods as sensitivity analyses. Our IVW analysis revealed that a one standard deviation increase in *ATG12* expression was associated with a significant 15% increase in the risk of intestinal obstruction (OR=1.15, 95% CI: 1.11-1.19, *P*=1.481e-13). Conversely, a one standard deviation increase in *CHRNB2* expression was associated with a significant 31% decrease in intestinal obstruction risk (OR=0.69, 95% CI: 0.62-0.78, *P*=2.754e-10). Similarly, a one standard deviation increase in *TNFRSF18* expression was associated with a significant 19% decrease in risk (OR=0.81, 95% CI: 0.78-0.84, *P*=7.146e-35). Finally, a one standard deviation increase in *MIAT* expression was associated with a significant 11% increase in risk (OR=1.11, 95% CI: 1.08-1.13, *P*=1.588e-20). The direction of effect estimates from other MR methods was generally consistent with the IVW results (Table 2).

**Table 2.**
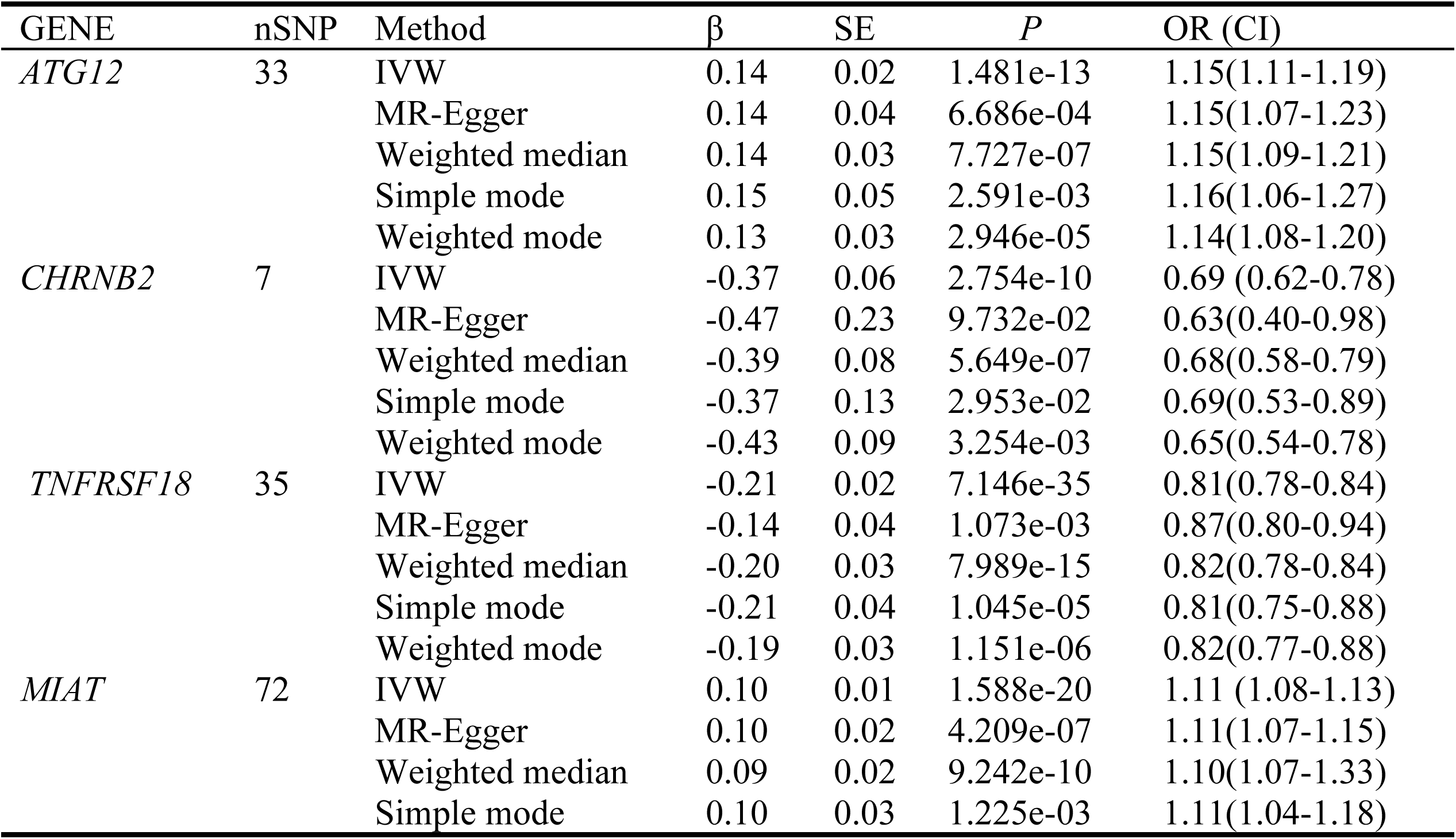
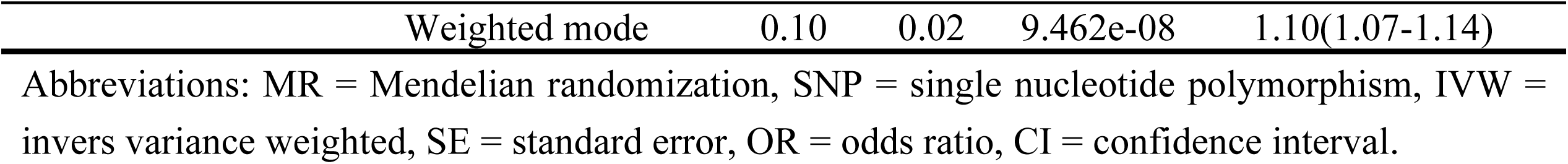
MR estimates for the effect of gene expression on the risk of intestinal obstruction.

To assess heterogeneity among the genetic instruments for each gene (Table 3), we employed Cochran’s Q test and Rücker’s Q test. These tests aim to evaluate whether the instruments influence the outcome through the same causal pathway. The results showed evidence of heterogeneity for the *MIAT* gene (Cochran’s Q = 96.72, *P* = 0.023), suggesting potential pleiotropy; while no heterogeneity was observed for the other genes (ATG12: Cochran’s Q = 23.85, *P*=0.850; *CHRNB2*: Cochran’s Q = 5.92, *P*=0.431; *TNFRSF18*: Cochran’s Q = 21.32, *P*=0.956). Horizontal pleiotropy was assessed using MR-Egger regression (Table 3). This method is used to examine whether genetic instruments are associated with the outcome through pathways other than the gene expression of interest. The MR-Egger regression intercept did not reach statistical significance for any of the genes (*P*>0.05), indicating no significant evidence of horizontal pleiotropy.

**Table 3.** Heterogeneity and horizontal pleiotropy assessment of the instrumental variables used in MR.

| GENE | Heterogeneity |  |  |  | Horizontal pleiotropy |  |  |
| --- | --- | --- | --- | --- | --- | --- | --- |
| | Rucker Q<br>MR-Egger | $P$ | Cochran Q<br>IVW | $P$ | Egger-intercept<br>MR-Egger | SE | $P$ |
| *ATG12* | 28.85 | 0.817 | 23.85 | 0.850 | 0.00 | 0.00 | 0.942 |
| *CHRNA2* | 5.68 | 0.337 | 5.92 | 0.431 | 0.01 | 0.03 | 0.667 |
| *TNFRSF18* | 17.51 | 0.988 | 21.32 | 0.956 | -0.02 | 0.01 | 0.060 |
| *MIAT* | 99.64 | 0.019 | 96.72 | 0.023 | -0.01 | 0.01 | 0.811 |

Abbreviations: IVW = inverse-variance weighted, MR = Mendelian randomization, SE = standard error.

Fig 2 visually presents forest plots for each gene, validating the primary findings of the IVW analysis. Forest plots in Fig 2A (*ATG12* gene) and Fig 2D (*MIAT* gene) demonstrate that both individual SNP instrument effect estimates and the IVW summary effect estimate are shifted to the right of the null effect line, visually indicating a positive association between increased *ATG12* and *MIAT* expression levels and an elevated risk of intestinal obstruction. Conversely, forest plots in Fig 2B (*CHRNB2* gene) and Fig 2C (*TNFRSF18* gene) display the opposite pattern, with both individual SNP effect estimates and the IVW summary effect estimate shifted to the left of the null effect line, visually indicating a negative association between increased *CHRNB2* and *TNFRSF18* expression levels and a reduced risk of intestinal obstruction.

**Fig 2.**
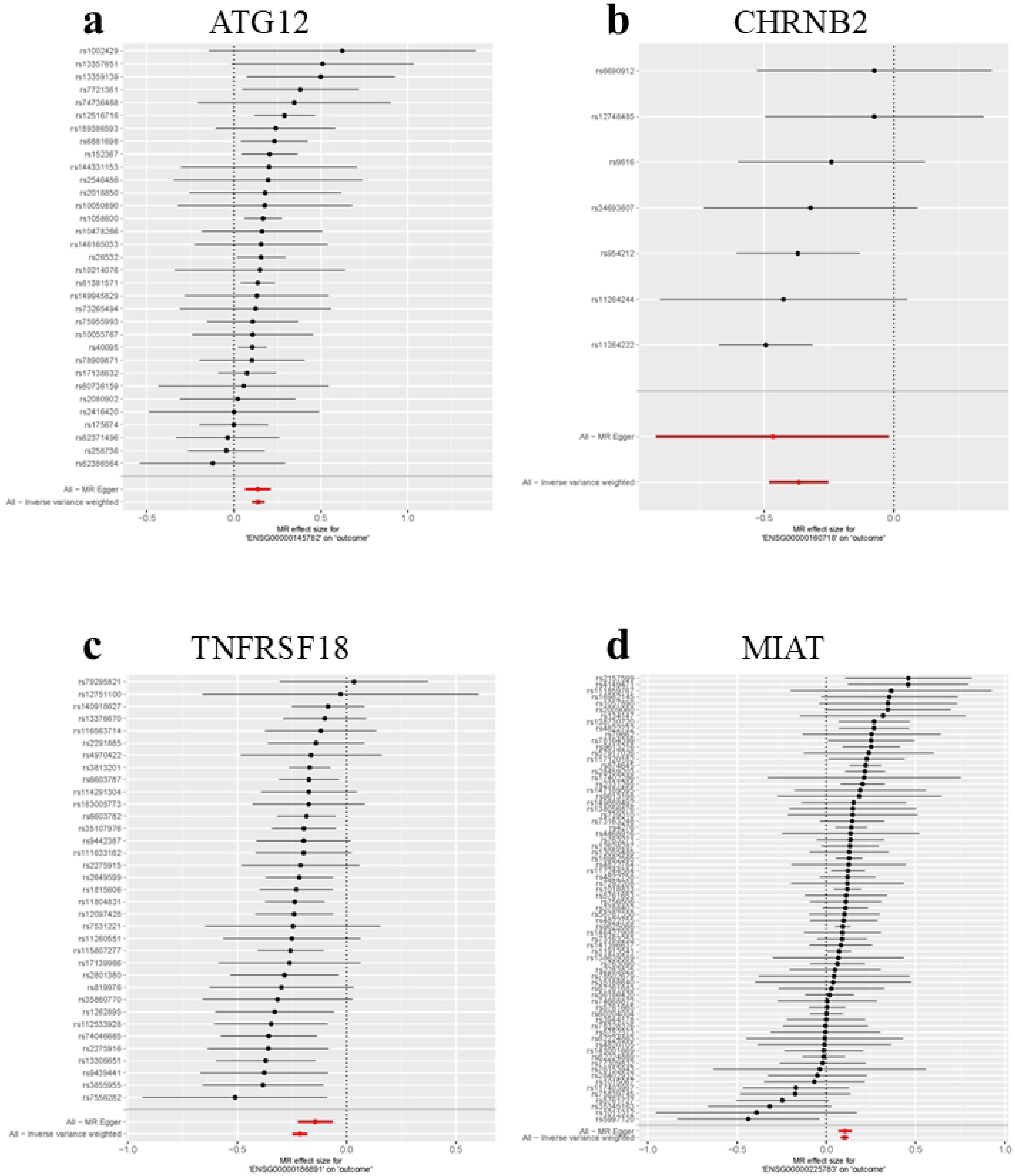
Causal effects of genetically predicted gene expression on intestinal obstruction risk: Forest plot results from Mendelian randomization analysis. Forest plots showing the Mendelian randomization (MR) estimates of the effect of genetically predicted expression levels of *ATG12* (A), *CHRNB2* (B), *TNFRSF18* (C), and *MIAT* (D) on the risk of intestinal obstruction. For each gene, the forest plot displays the effect estimate (black dot) and 95% confidence interval (horizontal line) for each genetic instrument (SNP). The red diamond represents the overall causal effect estimate and its 95% confidence interval, obtained using the inverse variance weighted (IVW) method.

Fig 3 visually demonstrates the relationship between SNP-exposure effects and SNP-outcome effects in the Mendelian randomization analysis. The linear trend observed in the scatter plots further validates the robustness of the IVW analysis results. The IVW regression line (blue) clearly illustrates the linear association pattern between SNP-exposure and SNP-outcome effects. Although the MR-Egger regression line (green) shows a slight deviation from the IVW line, their overall trends are largely consistent, and the MR-Egger intercept test P-value (P > 0.05) indicates no strong evidence of horizontal pleiotropy.

**Fig 3.**
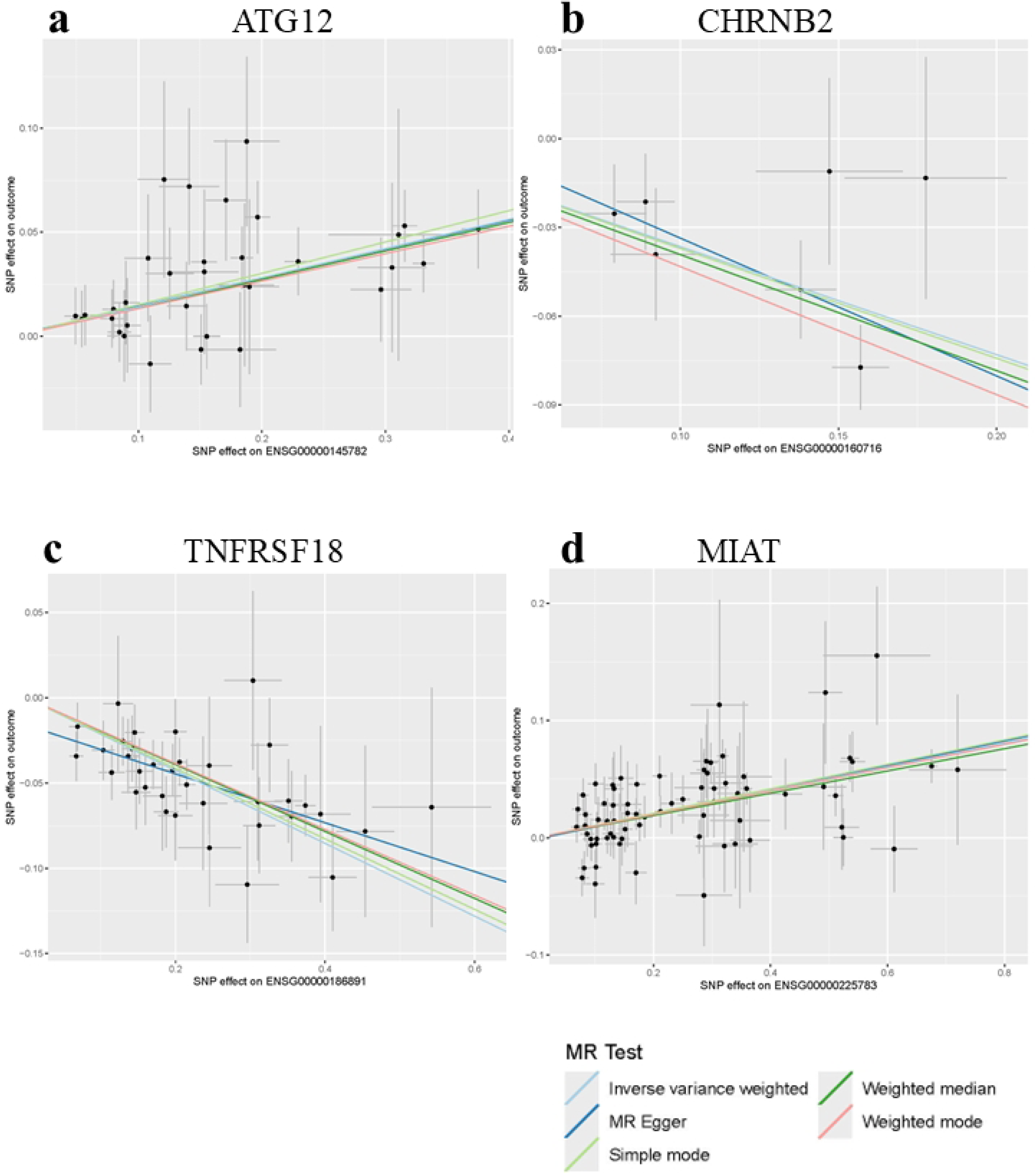
Scatter plots assessing linearity and pleiotropy in Mendelian randomization analysis of gene expression and intestinal obstruction. Scatter plots depicting the relationship between SNP-exposure effects and SNP-outcome effects for *ATG12* (A), *CHRNB2* (B), *TNFRSF18* (C), and *MIAT* (D) genes in the Mendelian randomization (MR) analysis. Each dot represents a SNP instrument. The blue line represents the Inverse Variance Weighted (IVW) regression line, the green line represents the MR-Egger regression line, the pink line represents the Weighted median regression line, the light blue line represents the Weighted mode regression line, and the yellow line represents the Simple mode regression line.

### Colocalization Analysis Results

To further assess whether the gene expression associations identified through MR share the same causal variants with intestinal obstruction GWAS signals, we performed colocalization analysis (COLOC). This analysis aims to estimate the posterior probability of a shared causal variant (PP4), with PP4 > 0.75 generally considered robust evidence for colocalization and these results are summarized in Table 4. Conversely, a PP3 > PP4 suggests that the GWAS association is likely driven by distinct causal variants rather than shared regulatory mechanisms with gene expression.

**Table 4.** Posterior Probabilities from Colocalization Analysis of Gene Expression and Intestinal Obstruction.

| Exposure | PP1 | PP2 | PP3 | PP4 |
| --- | --- | --- | --- | --- |
| *ATG12* | 3.28e-288 | 6.56e-01 | 2.23e-01 | 1.21e-01 |
| *CHRNA2* | 1.39e-63 | 6.58e-05 | 7.38e-02 | 9.26e-01 |
| *TNFRSF18* | 2.34e-56 | 5.71e-01 | 2.20e-01 | 2.09e-01 |
| *MIAT* | 7.95e-290 | 4.22e-03 | 1.58e-01 | 8.38e-01 |

Abbreviations: PP= Posterior Probabilities.

The colocalization analysis provided robust evidence for colocalization for *CHRNB2* (PP4 = 0.926) and *MIAT* (PP4 = 0.838), indicating that gene expression and GWAS signals are likely driven by the same underlying causal variants. These findings further strengthen the MR results, suggesting a potential causal role for *CHRNB2* and *MIAT* expression levels in the development of intestinal obstruction. In contrast, *ATG12* (PP4 = 0.121) and *TNFRSF18* (PP4 = 0.209) did not show strong evidence of colocalization, with PP3 exceeding PP4, implying that their GWAS associations may be more attributable to LD rather than genuine colocalization with eQTL regulation. Consequently, the MR findings for *ATG12* and *TNFRSF18* warrant further validation.

To visualize the colocalization signals for *CHRNB2* and *MIAT,* we generated colocalization comparison plots using the locuscomparer package (Supplementary Fig S1). Supplementary Fig S1 graphically illustrates the relationship between eQTL association signals (x-axis) and GWAS association signals (y-axis), providing a graphical representation of their colocalization. The results demonstrate that *CHRNB2* and *MIAT* exhibit strong overlap between eQTL and GWAS signals within the same genomic region, further reinforcing their biological relevance.

### SMR and HEIDI Validation Analysis Results

To further validate the MR findings, we conducted SMR analysis, and performed the HEIDI test to distinguish genuine colocalization from LD-driven associations (TABLE 5). SMR analysis confirmed significant associations for all four genes in the eQTLGen consortium database (*P*-SMR < 0.05), and also for *ATG12* and *MIAT* genes in the GTEx (intestinal tissue) database (*P*-SMR < 0.05). While data for *CHRNB2* and *TNFRSF18* genes were unavailable for SMR analysis in the GTEx (intestinal tissue) database (Table 5), and this data absence might be related to the relatively smaller sample size of the GTEx intestinal tissue database. HEIDI test results for all genes in the eQTLGen consortium database showed *P*-HEIDI > 0.01, supporting the hypothesis that the SMR signals are driven by colocalization rather than LD confounding. Acknowledging that COLOC analysis suggested potential LD influence for *ATG12* and *TNFRSF18* genes, we maintain necessary caution in interpreting the SMR findings.

**Table 5.**
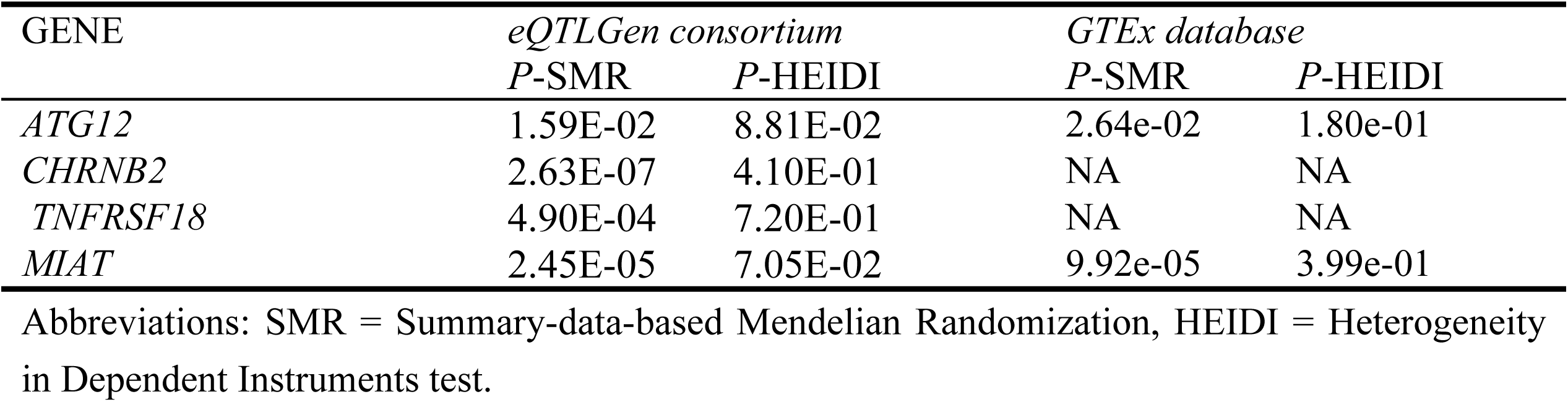
SMR and HEIDI validation analysis results for gene expression.

| GENE | *eQTLGen consortium* |  | *GTEx database* |  |
| --- | --- | --- | --- | --- |
|  | *P*-SMR | *P*-HEIDI | *P*-SMR | *P*-HEIDI |
| *ATG12* | 1.59E-02 | 8.81E-02 | 2.64e-02 | 1.80e-01 |
| *CHRNA2* | 2.63E-07 | 4.10E-01 | NA | NA |
| *TNFRSF18* | 4.90E-04 | 7.20E-01 | NA | NA |
| *MIAT* | 2.45E-05 | 7.05E-02 | 9.92e-05 | 3.99e-01 |

Abbreviations: SMR = Summary-data-based Mendelian Randomization, HEIDI = Heterogeneity in Dependent Instruments test.

## Discussion

This study systematically investigated the causal relationship between gene expression levels and the risk of intestinal obstruction using Mendelian randomization (MR). We focused on four candidate genes—*ATG12*, *CHRNB2*, *TNFRSF18*, and *MIAT* —which were selected based on their association with intestinal obstruction risk in our initial MR analysis. Leveraging data from the eQTLGen consortium, GTEx, and FinnGen studies, we sought to determine whether genetically predicted expression levels of these genes were causally related to intestinal obstruction risk. Our findings reveal potential causal associations between the expression levels of these four genes and the risk of intestinal obstruction. MR analysis indicated that increased expression of *ATG12* and *MIAT* was associated with increased risk, while increased expression of *CHRNB2* and *TNFRSF18* was associated with decreased risk. These findings were largely corroborated by subsequent colocalization and SMR validation analyses.

It is noteworthy that our colocalization analysis (COLOC) results presented a degree of divergence from our SMR analysis findings. Specifically, COLOC analysis suggested that the GWAS associations for *ATG12* and *TNFRSF18* might be attributed more to linkage disequilibrium (LD) confounding than to shared causal eQTL regulation, as partly evidenced by their lower PP4 values and relatively higher PP3 values. However, SMR analysis demonstrated significant associations between gene expression and intestinal obstruction risk for these two genes (*P*-SMR < 0.05). This discrepancy in findings may stem from several key factors. Firstly, inherent methodological differences exist between SMR and COLOC analyses. SMR analysis aims to estimate the overall causal effect of gene expression on disease risk, whereas COLOC analysis focuses on identifying shared causal variants. These methods diverge in their fundamental principles, underlying assumptions, and applicability, potentially contributing to inconsistent results [14]. For instance, SMR analysis assumes that instrumental variables influence disease risk solely through gene expression, while COLOC analysis posits a single causal variant influencing both gene expression and the disease outcome. Secondly, the complexity of linkage disequilibrium (LD) structure may also exert a significant influence on colocalization analysis. Complex LD relationships between causal and eQTL variants can potentially impede COLOC’s ability to precisely pinpoint shared causal variants[15]. If these variants reside within distinct LD blocks, or if multiple independent causal variants are present, COLOC may encounter challenges in effectively disentangling their individual effects. Thus, for *ATG12* and *TNFRSF18* genes, the inconsistency between COLOC and SMR results suggests that these associations may be confounded by LD rather than reflecting genuine colocalization (PP3 > PP4). Furthermore, discrepancies in sample size represent another potential contributing factor to the divergent findings. Our COLOC analysis utilized a smaller sample size compared to our SMR analysis. This disparity may have limited the statistical power of COLOC to detect true colocalization signals, while the larger sample size in the SMR analysis may have afforded greater statistical power to identify significant associations. Of particular note, the GTEx database, which exclusively comprises intestinal tissue samples (n=860) [8], is considerably smaller than the eQTLGen consortium database (n=31684). This difference in sample size likely accounts for the inability to detect eQTL variants for *CHRNB2* and *TNFRSF18* in GTEx, potentially impacting downstream analytical outcomes. The impact of sample size on statistical power is a crucial consideration in genetic studies[16].

Our findings elucidate potential causal links between the expression levels of *ATG12*, *CHRNB2*, *TNFRSF18*, and *MIAT*, and the risk of intestinal obstruction. *ATG12*, implicated in autophagy, a crucial cellular recycling process, may influence intestinal obstruction by modulating gut barrier integrity, immune responses, and inflammation [17]. *CHRNB2*, encoding a cholinergic receptor, regulates intestinal motility and secretion; its reduced expression may disrupt these physiological functions, thereby increasing the risk of obstruction [18]. *TNFRSF18*, an immune response regulator, likely impacts intestinal obstruction by modulating immune cell activity and cytokine release [19]. *MIAT*, a long non-coding RNA involved in gene regulation, may contribute to intestinal inflammation, potentially elevating obstruction risk[20]. It is important to acknowledge that our findings diverge from previous studies on *ATG12* in inflammatory bowel disease (IBD), which may be attributable to variations in study populations and methodologies [21]. Furthermore, our Mendelian randomization approach concentrates on elucidating causal relationships between gene expression and intestinal obstruction risk, whereas other studies may investigate gene function *in the context of* inflammatory bowel disease; these differences in research focus may also contribute to the observed discrepancies.

While this study elucidates potential causal links between gene expression and intestinal obstruction risk, several limitations warrant consideration. Primarily, the predominantly European ancestry of the study population may limit the broad generalizability of our findings.

Methodologically, although causal relationships are established, exploration of underlying regulatory mechanisms remains limited; the use of whole blood eQTL data restricts comprehensive interpretation of intestinal tissue-specific regulation; and the absence of direct functional validation necessitates further investigation. Statistically, SMR analysis power is contingent on eQTL sample size, and the single causal variant assumption of COLOC analysis may oversimplify the complexity of certain loci. Furthermore, residual confounding from unmeasured factors, such as dietary habits and gut microbiota composition, cannot be entirely discounted. Addressing these limitations in future research will enhance the understanding of the genetic and molecular mechanisms underlying intestinal obstruction and improve the reliability of Mendelian randomization-based findings.

## Conclusions

This study furnishes genetic evidence supporting the causal roles of *CHRNB2* and *MIAT* in intestinal obstruction. While SMR analysis suggests potential associations for *ATG12* and*TNFRSF18*, further validation is warranted for these genes due to potential LD confounding.

These findings offer novel insights into the pathogenesis of intestinal obstruction and identify potential therapeutic targets for its prevention, diagnosis, and treatment.

## Acknowledgements

We thank Dr. Ni wentao (Peking University People’s Hospital, Peking, China) for helping in this revision.

## Data Availability

Data in the article can be obtained from *eQTLGen consortium* (https://www.eqtlgen.org/), *FinnGen consortium* (https://r12.finngen.fi) a*nd GTEx database* (https://gtexportal.org).

## Additional Information

This research received no specific grant from any funding agency in the public, commercial or not-for-profit sectors. No potential conflict of interest was reported by the author(s).

## Supporting information

**S1 Fig.**
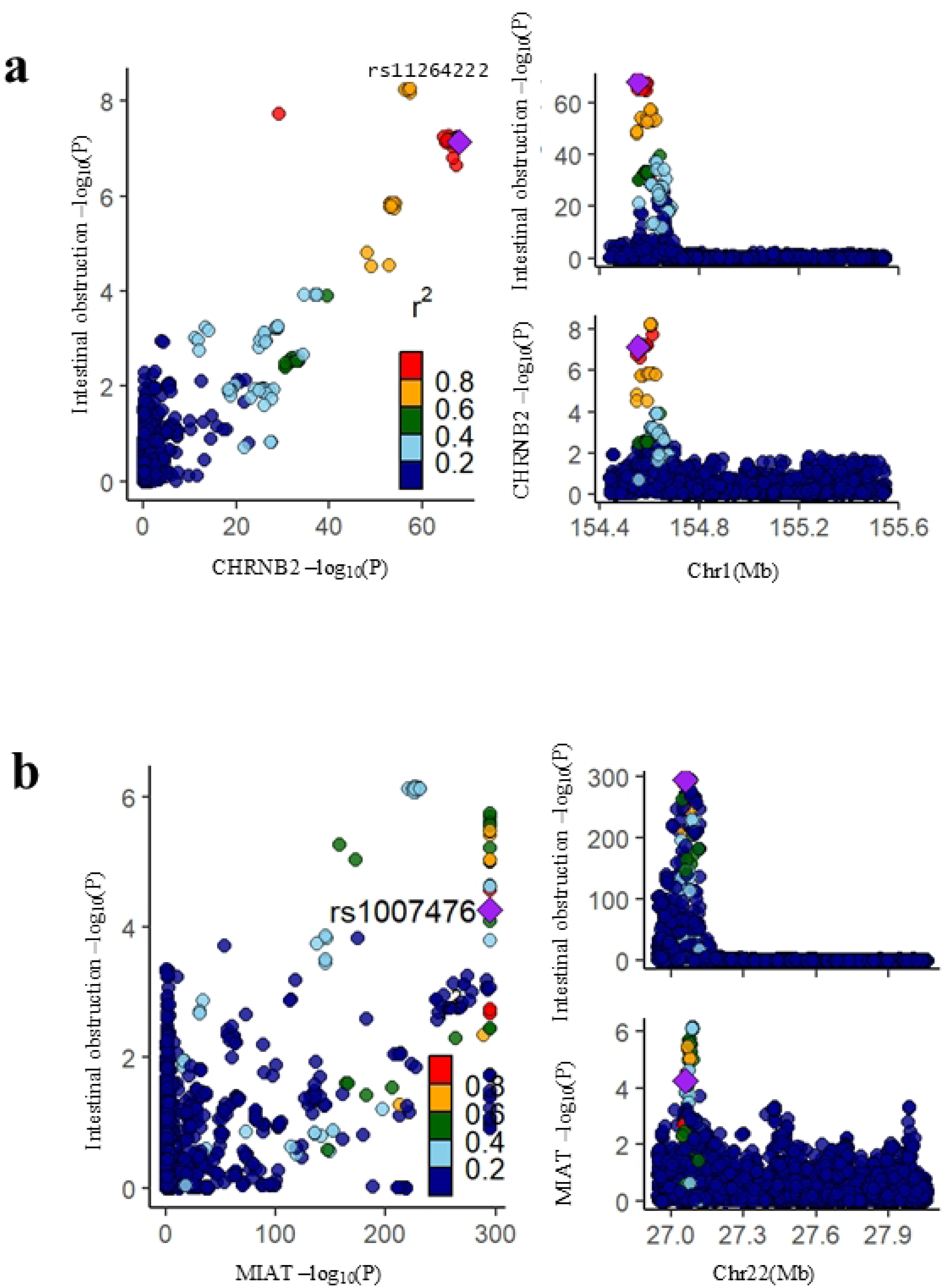
Colocalization comparison plots for *CHRNB2* and *MIAT* genes. This Fig displays the colocalization analysis results for *CHRNB2* (A) and *MIAT* (B) genes, generated using the locuscomparer package. Left panels: Scatter plots. The X-axis represents eQTL association signals (−log10(P)), and the Y-axis represents intestinal obstruction GWAS association signals (−log10(P)). Each point represents a SNP, colored by linkage disequilibrium (LD, r²) with the lead SNP. Right panels: Regional association plots. The X-axis represents chromosomal position (Mb), and the Y-axis represents intestinal obstruction GWAS association signals (−log10(P)). The plots show the distribution of GWAS signals across the gene region, with the lead SNP marked as a purple diamond (rs11264222 for *CHRNB2*, rs1007476 for *MIAT)*.

**S1 Table Final genetic instrumental variables (IVs) selected from eQTLGen for Mendelian randomization analysis evaluating the causal association of gene expression with intestinal obstruction.** This table presents the final set of genetic IVs selected from the eQTLGen Consortium (Release 2) *cis*-eQTL dataset using predefined filtering criteria (FDR < 0.05, LD R² < 0.1) to ensure the independence and statistical robustness of the IVs. β, SE, and P-values represent the association results between the IVs and gene expression.

Abbreviations: IVs = instrumental variables, MR = Mendelian randomization, SNP = single-nucleotide polymorphism, IVW = inverse-variance weighted, SE = standard error, OR = odds ratio, CI = confidence interval.

**S2 Table List of candidates *cis*-eQTL variants from eQTLGen for Mendelian randomization analysis of intestinal obstruction.** This table provides a comprehensive list of candidates *cis*-eQTL variants, initially extracted from the eQTLGen Consortium (Release 2) dataset, considered for use as instrumental variables in the Mendelian randomization analysis evaluating the causal association between gene expression and intestinal obstruction risk. Two-sample Mendelian randomization analysis was performed using the inverse-variance weighted (IVW) method to estimate the effect size of gene expression on intestinal obstruction risk. To account for multiple testing, statistical significance was determined using a Bonferroni-corrected P-value threshold of *P* < 3.14 × 10⁻⁷ (α=0.05 / 159,339).

Abbreviations: eQTL = expression quantitative trait locus, GWAS = genome-wide association study, MR = Mendelian randomization, IVW = inverse-variance weighted, FDR = false discovery rate, LD = linkage disequilibrium.

